# Direct utility of natural history data in analysis of clinical trials: Propensity match-based analysis of Omaveloxolone in Friedreich ataxia using the FA-COMS dataset

**DOI:** 10.1101/2022.08.12.22278684

**Authors:** David R Lynch, Angie Goldsberry, Christian Rummey, Jennifer Farmer, Sylvia Boesch, Martin B. Delatycki, Paola Giunti, J. Chad Hoyle, Caterina Mariotti, Katherine D. Mathews, Wolfgang Nachbauer, Susan Perlman, S.H. Subramony, George Wilmot, Theresa Zesiewicz, Lisa Weissfeld, Colin Meyer

## Abstract

**Rationale:** The natural history of Friedreich Ataxia (FRDA) is being investigated in a multi-center longitudinal study designated the Friedreich Ataxia Clinical Outcome Measures Study (FA-COMS). To understand the utility of this natural history dataset in analysis of clinical trials, we performed a propensity-matched comparison of the data from the open-label MOXIe Extension (omaveloxolone) with that from FA-COMS.

**Methods:** All MOXIe Extension patients who had at least one post-baseline assessment were matched to FA-COMS patients using logistic regression to estimate propensity scores based on multiple covariates: sex, baseline age, age of onset, baseline modified Friedreich Ataxia Rating scale (mFARS) score, and baseline gait score. Selection of covariates was based on clinical relevance (i.e., factors considered prognostic for disease progression) and availability. The change from baseline in mFARS at Year 3 for the MOXIe Extension patients compared to the matched FA-COMS patients was analyzed as the primary efficacy endpoint using mixed model repeated measures analysis.

**Results:** Data from the MOXIe Extension show that omaveloxolone provides persistent benefit over three years when compared to an untreated, rigorously matched cohort from FA-COMS. At each year, and in all analysis populations, patients in the MOXIe Extension experienced a smaller change from baseline in mFARS score than the matched FA-COMS patients. In the Primary Pooled Population (136 patients in each group) by Year 3, patients in the FA-COMS matched set progressed 6.6 points whereas patients treated with omaveloxolone in MOXIe Extension progressed 3 points (difference =-3.6; nominal p value =0.0001). Thus, progression in mFARS was slowed by 55% with omaveloxolone treatment.

**Conclusions:** These results suggest a clinically meaningful slowing of FRDA progression with omaveloxolone, and consequently details how propensity-matched analysis contributes to the understanding of the effects of therapeutic agents. This demonstrates the direct value of natural history studies in the evaluation of clinical trials.

---

Friedreich ataxia (FRDA) is an autosomal recessive neurodegenerative disorder resulting from deficiency of the protein frataxin [1–3]. This deficiency leads to decreased ATP production, abnormalities of oxidative phosphorylation, and a diminished antioxidant response [1–6]. Suppression of activity of the transcription factor Nrf2, which has been documented in FRDA patients and animal models, contributes to impairment of mitochondrial energy production and oxidative damage [7–10]. These cellular events lead to the clinical phenotype of progressive ataxia, dysarthria, sensory loss, dyscoordination, and cardiomyopathy. While neurological dysfunction is present in all subjects and is relentlessly progressive, the speed of progression is heterogeneous based on genetic severity and likely other factors [11–13].

The clinical features of FRDA have become better understood based on data from large ongoing natural history studies. Beginning in 2003, the Friedreich Ataxia Clinical Outcome Measures Study (FA-COMS) follows more than 1250 participants enrolled at 15 participating clinical sites [12–15]. It (along with a parallel study in Europe called EFACTS) [11] has mapped the disease course, helped develop outcome measures and biomarkers, and generated more than 30 scientific publications, thus establishing the infrastructure for conducting more than 10 interventional FRDA clinical trials [16–17]. FA-COMS has been primarily focused on measuring the neurological symptoms of FRDA, as well as cardiac disease, vision changes, scoliosis, diabetes and other medical conditions, and medications. The study has led to the development and validation of the Friedreich Ataxia Rating Scale (FARS), a scale including neurologic signs and functional assessments reflecting specific neural substrates affected in FRDA [14-15,18]. Removing items of limited functional significance (such as peripheral nerve elements) improved the measure, creating the modified FARS or mFARS, now used as the primary outcome measure in multiple clinical trials. This includes the MOXIe study, a randomized, double-blind trial that demonstrated a benefit of omaveloxolone in FRDA [19–20].

In addition to characterizing the disease and developing outcome measures, natural history studies can also provide control populations for other clinical studies, including intervention studies. This is most important in situations in which a true placebo is not available (surgical interventions), or study subjects are difficult to recruit as is the case for rare diseases such as FRDA. Although clinical trials and natural history studies do not provide identical study populations, a variety of approaches can be used to minimize bias and allow a systematic assessment of therapeutic response. The size of the FA-COMS database makes a propensity score analysis with optimal 1:1 matching strategy feasible. Therefore, the comparability of the two studies was evaluated to determine the suitability of the FA-COMS dataset as a source of relevant external control data for omaveloxolone in the MOXIe Extension study. In the present study, we report a propensity-matched analysis between MOXIe Extension data and FA-COMS natural history data to assess the value of this natural history study in understanding the effects of omaveloxolone in FRDA.

## METHODS

### FA-COMS

FA-COMS (NCT03090789), the largest, global, multi-center, prospective natural history study in FRDA, began enrolling patients in 2003 and is continuously enrolling, with more than 1,000 patients to date [13–17]. Patients are evaluated annually on FARS/mFARS, other neurologic outcomes, and quality-of-life assessments. All sites receive training on the protocol and collection of procedures as well as on data entry into standardized case report forms. Additionally, the study principal investigators meet every 3-6 months to review the overall conduct of the study, data analysis, results and findings, publications, and study-related issues. All subjects provided written informed consent, and the study was approved by local IRBs.

### MOXIe study Extension

The time period for FA-COMS overlaps with the MOXIe study (NCT02255435), which consists of 3 parts (Part 1, Part 2, and Extension); the first patient enrolled in the Part 2 portion in October 2017, the last visit occurred in October 2019, and the Extension study remains ongoing. All subjects provided written informed consent, and the study was approved by local IRBs. The ongoing MOXIe Extension assesses long-term safety and tolerability of omaveloxolone in patients with FRDA who completed MOXIe Part 1 or Part 2, both of which were placebo-controlled [19–20]. Patients and investigators remain blinded to their preceding study treatment throughout the Extension. All patients enrolled in the Extension receive open-label omaveloxolone (150 mg) once daily. Efficacy assessments including the mFARS are conducted at baseline (Extension Day 1) and then every 24 weeks. <u>Primary Endpoint</u>. The mFARS is a zero-to 99-point scale comprised of subsections A, B, C, and E of the FARS. A higher score on the mFARS examination signifies more severe physical impairment, and a reduction in score signifies functional improvement [15, 21]. For the present study, the primary efficacy endpoint was the change in mFARS score from baseline at Year 3 while secondary endpoints were the change in mFARS score from baseline at Years 1 and 2.

#### Ethics Statement

All subjects provided written informed consent. The protocol was approved by the IRB/ Ethics committees at all Institutions: The Children’s Hospital of Philadelphia (lead site), UCLA, University of Iowa, Emory, University of South Florida, University College London, University of Florida, University of Melbourne/Murdoch Research Institute (Australia), Neurological Institute Carlo Besta (Italy), Medical University of Innsbruck (Austria), and Ohio State University.

### Statistical plan

The methodology for these post-hoc analyses, including the determination of covariates used for determining propensity scores for matching, was developed with several FRDA experts, including the lead author (DRL), the FA-COMS statistician (CR), representatives from FARA (JF), statisticians at Reata (AG), and external statisticians from WCG-Statistics Collaborative (LR). This study uses data from a 24 March 2022 interim database lock for the ongoing MOXIe Extension. The FA-COMS dataset was current as of 24 March 2021 when it was obtained from the Critical Path Institute (c-Path)(https://c-path.org/programs/dcc/projects/friedreichs-ataxia/). FA-COMS data is publicly available through the Critical Path Institute as part of the Friedreich Ataxia Integrated Clinical Database (FA-ICD).

In the analysis, data were compared between MOXIe Extension patients (treatment) and matched FA-COMS natural history patients (external control). The study populations were derived from the full MOXIe Extension dataset (n=149) and the full FA-COMS dataset (n=810) (**Figure 1A**). For inclusion in each of the study populations, patients must have had a baseline mFARS, at least one post-baseline mFARS within 3 years after baseline, and values for all propensity score model covariates (i.e., sex, baseline mFARS score, age at baseline, age of FRDA onset, baseline gait score). All patients were included, regardless of pes cavus status. Three study populations were defined: the Moxie Extension population, the natural history (NH) population, and the sensitivity NH population (SNH). The SNH was defined as the subset of patients from the NH population who had a baseline mFARS score within the range observed at baseline in the MOXIe Extension (mFARS 8 to 74) and an age at baseline within the MOXIe Extension baseline range (16 -41 years). These were chosen as additional selection criteria because, on a population level, age is the best predictor of mFARS progression rate [13], and baseline mFARS score best controls for ceiling effects in the mFARS scale [14].

**Figure 1:**
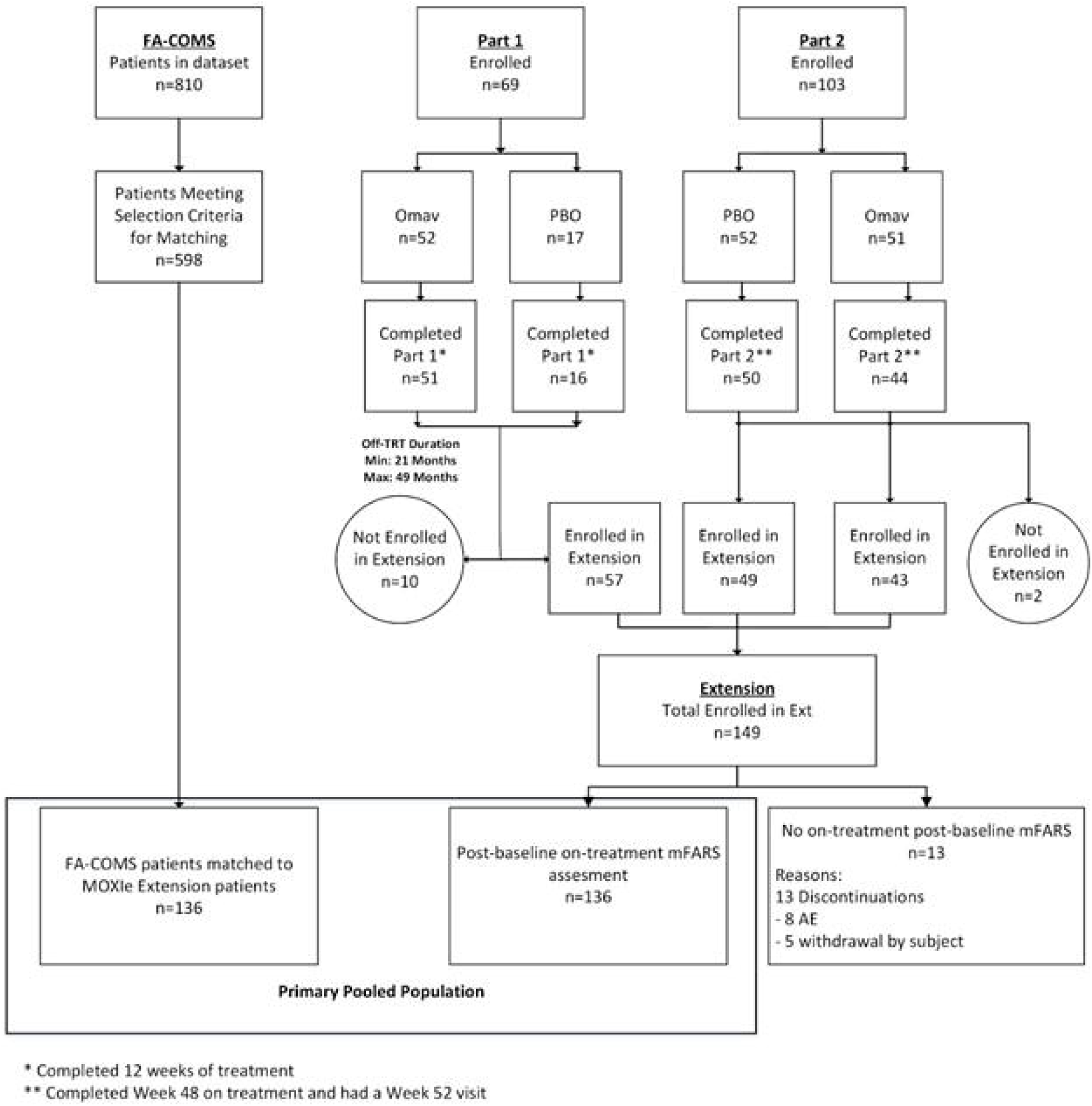
Diagram of Study populations (A), Moxie selection sets (B), and matching process (C) **A. Study Populations** Populations were selected from the FA-COMS and MOXIe Extension studies/ All study populations require at least one post-baseline mFARS within 3 years after baseline, and values for all propensity score model covariates. For the sensitivity natural history population, Baseline mFARS and age were within the range observed in MOXIe Extension patients at baseline. **B. MOXIe Extension Sets of Patients** From the Extension participants, subjects were further classified as to whether they were on sustained omaveloxolone before starting the Extension. **C. Primary Analysis Populations** The propensity matching process was carried out separately for the 3 analysis sets.

### Analysis Populations

The MOXIe Extension population (**Figure 1B**) can be divided into 2 sets of patients based on their prior treatment status: (1) patients considered treatment-naïve, and (2) patients continuing treatment from their prior study. The first patient was enrolled in the Extension after Part 2 enrollment was complete. For MOXIe Part1 patients, this resulted in a minimum 21-month off-treatment period prior to enrolling in the MOXIe Extension (**Figure 2**). Due to this long off-treatment period and the short treatment duration in Part 1 (12 weeks), patients from Part 1 were considered treatment-naïve upon entry into the Extension and included in the placebo-omaveloxolone (Placebo-Omav) group. Patients who received placebo in Part 2 were also considered treatment-naïve and included in the Placebo-Omav group. Only those patients who received omaveloxolone in Part 2 and continued treatment in the Extension were included in the omaveloxolone-omaveloxolone (Omav-Omav) group.

**Figure 2:**
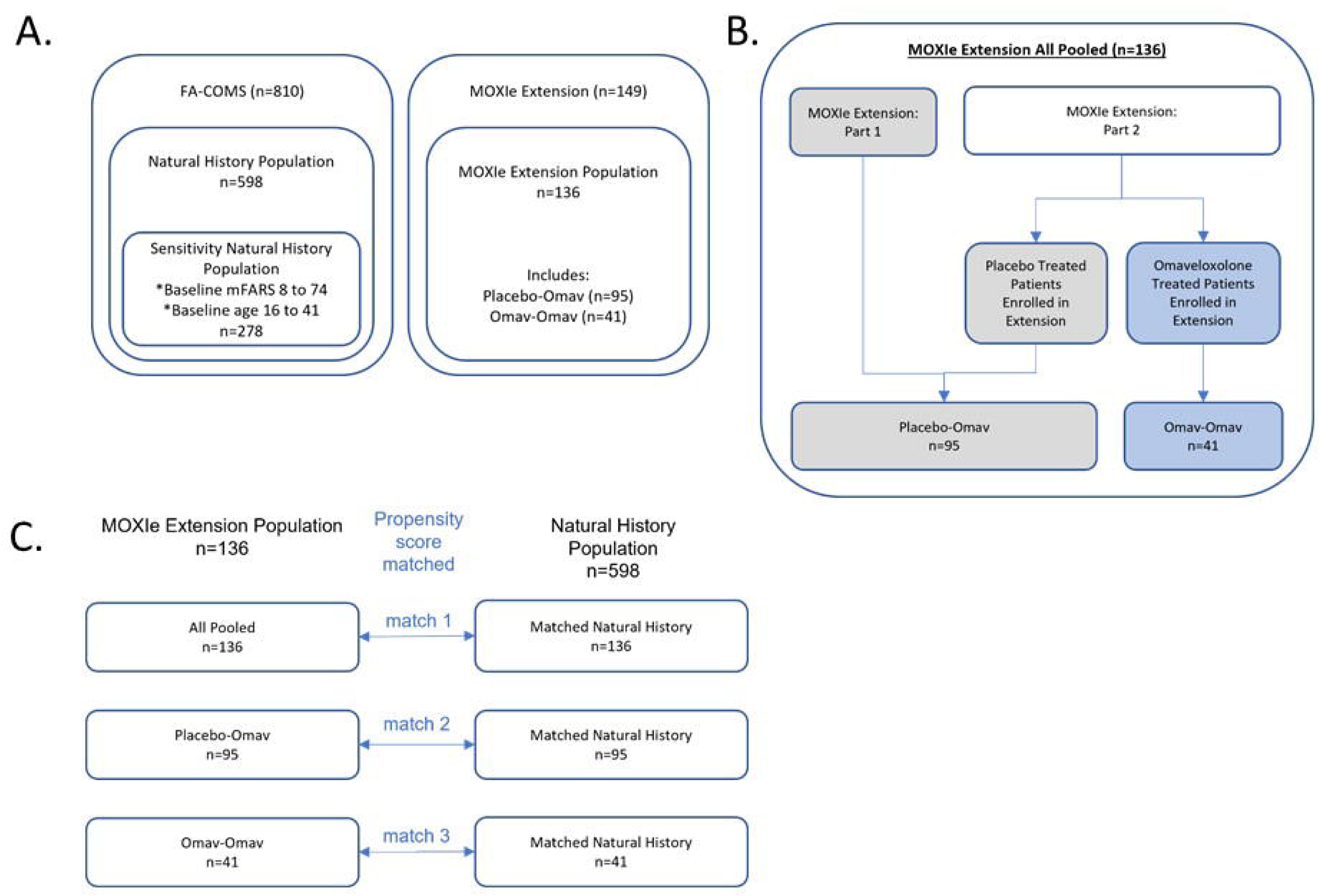
CONSORT diagram of entire study.

Each of the analysis populations was based on a new propensity score match. The primary analysis populations (i.e., Primary Pooled, Primary Placebo-Omav, Primary Omav-Omav) were based on matches with the NH population (**Figure 1C**). The sensitivity analysis populations (i.e., Sensitivity Pooled, Sensitivity Placebo-Omav, Sensitivity Omav-Omav) were based on matches with the SNH population.

### Propensity score matching

The goal of propensity score matching is to create comparable groups through the estimation of the propensity score, defined as the probability that a patient received omaveloxolone (i.e., enrolled in MOXIe Extension) given a set of covariates. Propensity score matching mimics some characteristics of a randomized study. The observed covariates used for determining propensity scores are controlled for in patients having the same propensity score. Therefore, differences between the MOXIe Extension and FA-COMS groups should be accounted for and are likely not a result of observed covariates.

The propensity score is a linear combination of the covariates requiring that patients have a similar propensity score rather than a caliper match on a group of covariates. Computation of the propensity score was coupled with diagnostics to assess the adequacy of matching techniques used in the analysis. The matching was carried out as optimal 1:1 matching without replacement.

Several assumptions were made when creating the analysis populations for the proposed design and propensity score computation. These assumptions include:

- Strongly ignorable treatment assignment [22]: The treatment assignment must be independent of the change from baseline in mFARS score over time given the covariates used in the analysis. There is a positive probability of being in the omaveloxolone or the FA-COMS population, that is the propensity score estimated from the logistic regression model must be strictly greater than 0 and less than 1.
- Stable-unit treatment value assumption [23–24]: The outcomes of one individual are not affected by the group assignment of another.

These assumptions were met in this approach using propensity scores.

### Computation of the propensity score

The propensity score was estimated using logistic regression with covariates. The criteria for determining model fit differ from those for standard logistic regression analysis, as the goal of propensity score analysis is to balance key covariates across the MOXIe Extension patients and control patients, not to estimate a treatment effect. Omission of covariates potentially related to the outcome could increase the bias, arguing for a strategy of including more, rather than fewer covariates in the model.

Factors established as prognostic and available in both FA-COMS and MOXIe studies were selected as covariates for the logistic regression model used for determining propensity scores (**Table 1**)[21, 25]. Some factors, such as GAA1 repeat (the shorter of the two *FXN* intron 1 GAA repeats), may be prognostic but were not available for all patients. Notably, the presence of pes cavus was not a matching criterion for the FA-COMS external cohort as it was not systematically evaluated in the same manner as in the MOXIe Extension or available for all patients.

**Table 1:**
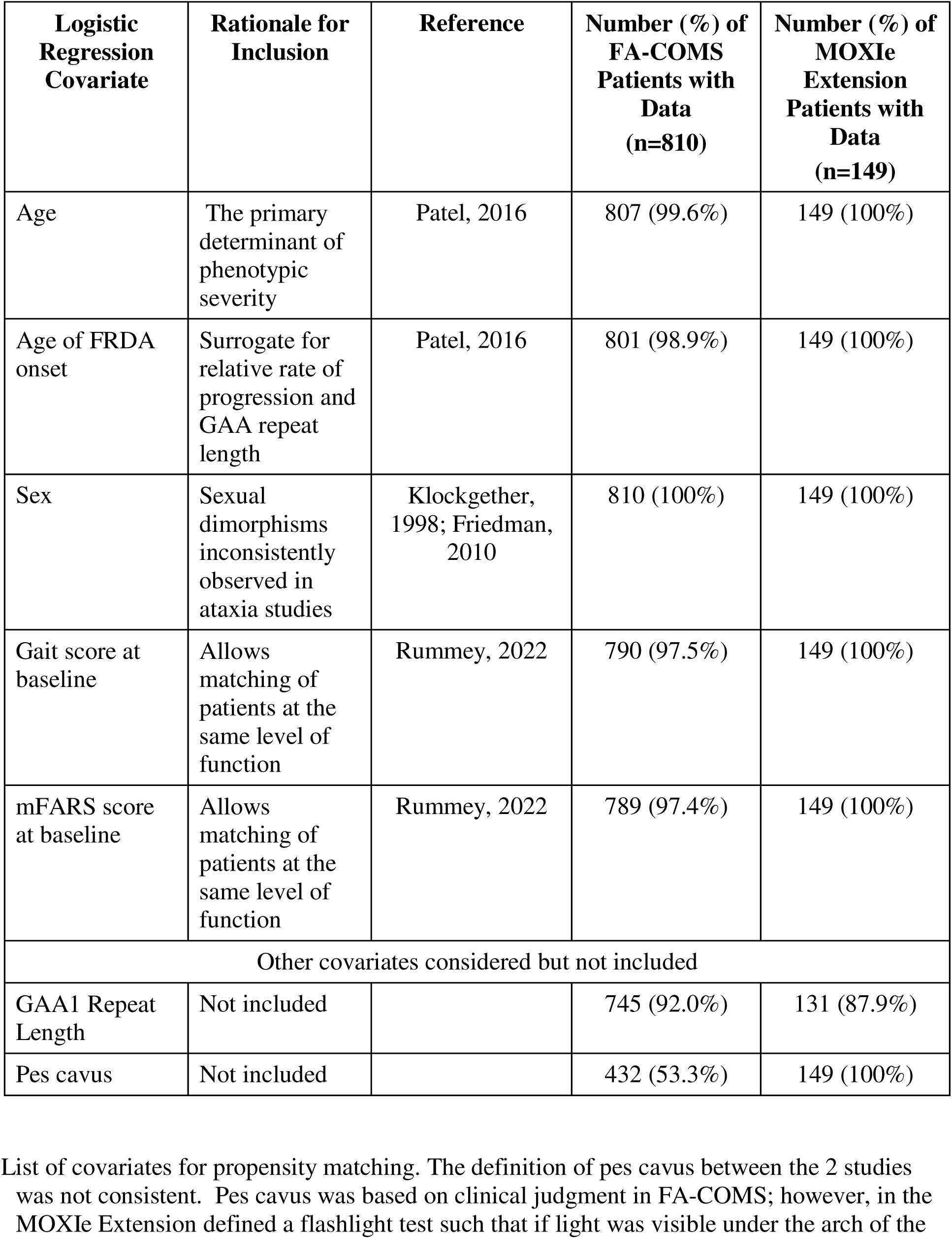
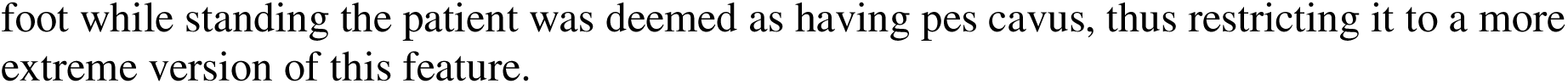
Covariates Used in propensity score matching

| **Logistic Regression Covariate** | **Rationale for Inclusion** | **Reference** | **Number (%) of FA-COMS Patients with Data (n=810)** | **Number (%) of MOXIe Extension Patients with Data (n=149)** |
| --- | --- | --- | --- | --- |
| Age | The primary determinant of phenotypic severity | Patel, 2016 | 807 (99.6%) | 149 (100%) |
| Age of FRDA onset | Surrogate for relative rate of progression and GAA repeat length | Patel, 2016 | 801 (98.9%) | 149 (100%) |
| Sex | Sexual dimorphisms inconsistently observed in ataxia studies | Klockgether, 1998; Friedman, 2010 | 810 (100%) | 149 (100%) |
| Gait score at baseline | Allows matching of patients at the same level of function | Rummey, 2022 | 790 (97.5%) | 149 (100%) |
| mFARS score at baseline | Allows matching of patients at the same level of function | Rummey, 2022 | 789 (97.4%) | 149 (100%) |
| **Other covariates considered but not included** |  |  |  |  |
| GAA1 Repeat Length | Not included |  | 745 (92.0%) | 131 (87.9%) |
| Pes cavus | Not included |  | 432 (53.3%) | 149 (100%) |

List of covariates for propensity matching. The definition of pes cavus between the 2 studies was not consistent. Pes cavus was based on clinical judgment in FA-COMS; however, in the MOXIe Extension defined a flashlight test such that if light was visible under the arch of the
foot while standing the patient was deemed as having pes cavus, thus restricting it to a more extreme version of this feature.

The logistic regression modeling, matching, and assessment of the quality of the matches were executed using PROC PSMATCH in SAS with additional diagnostics computed separately. The following options within PSMATCH were used:

- The region used was to be that of the common support (REGION = CS).
- Optimal matching was used to create the matches.
- Matching for sex was exact, i.e., females were only matched to females and males were only matched to males.
- The distance between matches was measured on the logit of the propensity score.
- The caliper (the specified distance between observations for declaring a “match”) for matching was set to missing and no weighting was used.
- The assessment for the quality of matches was conducted for the logit of the propensity score, the propensity score, and all covariates in the model.

The estimated propensity score *ê*, for the *i*^th^ patient was computed as follows:

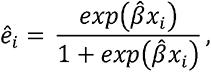

where 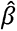 denotes the estimate of (*β* obtained from a logistic model, where the probability that a patient is a member of the treated group given a covariate vector x, written as P(Y=1|*x*), was defined as follows:

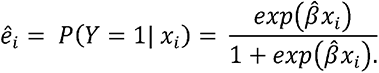

Note that *ê_i_*, is a probability with 0 ≤ *ê_i_*, ≤ 1. The analysis used the estimated linear propensity score *g_i_*, which was calculated as 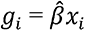.

### Creation of the analysis populations from the propensity analysis

After computation of the initial propensity score for males and females, optimal matching was used to create the matched population that was used for the analysis. Diagnostics were then used to assess the similarity of the two groups and whether the propensity score model was adequately specified.

### Efficacy Analysis Considerations

#### Definition of baseline

Baseline values were defined as the last non-missing assessment prior to the first study drug administration in MOXIe Extension. For FA-COMS patients, baseline was defined as having a record marked VISIT = Day 1. As an example, if a patient had no recorded FARS assessment at the baseline visit, but they did have a FARS assessment at Year 2, the Year 2 assessment was not considered as baseline and the patient was not included in the analysis as they did not have a baseline assessment.

#### Primary Efficacy analyses

The change from baseline in mFARS at Year 3 was analyzed using a mixed model repeated measures (MMRM) model that included treatment group, baseline mFARS, visit, and interaction terms for visit-by-baseline and treatment group-by-visit as covariates. The model was fit using restricted maximum likelihood with a Toeplitz covariance structure (assuming different variances at each time point and that measurements taken closer together in time are more highly correlated than those taken farther apart).

#### Visit schedule

Annual visits for the MMRM analysis were defined to align with the FA-COMS assessment schedule. The mFARS assessment collected closest to 1, 2, and 3 years after baseline was used for each of the annual assessments.

#### Secondary and sensitivity analyses

The secondary endpoints were analyzed using the same MMRM model as for the primary outcome of change from baseline in mFARS at Year 3. For sensitivity analyses, the analyses above were repeated for each sensitivity population.

## RESULTS

### Patients

Overall, 149 patients were enrolled in the MOXIe Extension, 57 patients from Part 1 and 92 from Part 2 (**Figure 2**). Of these, 136 had a post-baseline mFARS assessment while on treatment and were included in the propensity-matched analysis for the Primary Pooled population; the other 13 were excluded from further study. For the 136 patients in the Primary Pooled Population, the median treatment duration in MOXIe Extension (exclusive of treatment duration in Part 1 or Part 2) was 2.76 years, with a maximum of 3.4 years and a minimum of 0.5 years as of 24 March 2022 (**Table 2**).

**Table 2:** Follow-Up/Exposure Duration (Primary Pooled Population)

| Statistic | Matched FA-COMS Follow-Up<br>(N=136) | MOXIe Extension Exposure<br>(N=136) |
| --- | --- | --- |
|  | Years | Years |
| Mean (SD) | 2.54 (0.786) | 2.60 (0.524) |
| Median | 2.92 | 2.76 |
| Min, Max | 0.6, 3.5 | 0.5, 3.4 |

Follow-up duration in the natural history study is reported for the matched FA-COMS patients, and exposure to omaveloxolone is reported for MOXIe Extension patients.

The FA-COMS dataset received from C-Path included 810 patients who consented to have their data shared outside of the core FA-COMS study. Of these 810 patients, 598 met the criteria for inclusion in the NH study population, and 278 patients met criteria for inclusion in the SNH study population. These were included as potential matches to patients in the MOXIe Extension. The FA-COMS external cohort that was the matched set for the MOXIe Extension patients in the Primary Pooled population consisted of 136 patients. These patients had a median follow-up duration in the ongoing FA-COMS natural history study of 2.92 years, with a maximum of 3.5 years and a minimum of 0.6 years (**Table 2**). In total, the Primary Pooled population included 272 patients (136 from the MOXIe Extension and 136 matched patients from FA-COMS).

### Demographics and baseline characteristics

Covariates used for determining the propensity scores were well-balanced between the groups (**Table 3**) as were other demographics and baseline characteristics (**Table 4**). Slight differences observed in GAA1 and GAA2 (the longer of the two *FXN* GAA intron 1 repeats) repeat length were not clinically meaningful based on ceiling effects of the GAA1 length [26–28]. Demographics and baseline characteristics for the Primary Placebo-Omav and Primary Omav-Omav populations were generally well-balanced between the groups in both populations, although differences as in the Primary Pooled population in GAA1 and GAA2 repeat length were observed (**Supplementary Tables 1,2** respectively). Similarly, demographics and baseline characteristics were well-balanced in the Sensitivity Pooled, Sensitivity Placebo-Omav, and Sensitivity Omav-Omav populations (**Supplementary Tables 3,4,5**). In addition, no systematic differences were noted in concomitant medications between the different groups (data not shown).

**Table 3:** Demographics and Baseline Characteristics Used as Covariates for Propensity Score Calculation (Primary Pooled Population)

| Characteristic | Statistic | Matched FA-COMS | MOXIeExtension |
| --- | --- | --- | --- |
| Age (years) | n | 136 | 136 |
|  | Mean (SD) | 26.2 (13.72) | 26.6 (7.26) |
| Age at FRDA Onset | n | 136 | 136 |
|  | Mean (SD) | 15.2 (10.48) | 15.5 (5.30) |
| Sex (n [%]) | n | 136 | 136 |
|  | Female | 70 (51.5%) | 70 (51.5%) |
|  | Male | 66 (48.5%) | 66 (48.5%) |
| mFARS | n | 136 | 136 |
|  | Mean (SD) | 41.0 (16.10) | 42.2 (12.60) |
| Gait (Assessment #7 in FARS Section E [Upright Stability]) | n | 136 | 136 |
|  | Mean (SD) | 2.7 (1.69) | 2.8 (1.36) |

**Table 4:** Other Demographics and Baseline Characteristics (Primary Pooled Population)

| Characteristic | Statistic | Matched FA-COMS | MOXIe Extension |
| --- | --- | --- | --- |
| Ethnicity (n [%]) | n | 136 | 136 |
|  | Hispanic or Latino | 6 (4.4%) | 6 (4.4%) |
|  | Not Hispanic or Latino | 129 (94.9%) | 130 (95.6%) |
|  | Not reported | 1 (0.7%) | 0 |
| Race (n [%]) | n | 130 | 136 |
|  | White | 125 (96.2%) | 133 (97.8%) |
|  | Non-White | 5 (3.8%) | 3 (2.2%) |
| Height (cm) | n | 89 | 136 |
|  | Mean (SD) | 165.101 (14.6676) | 169.305 (10.4216) |
| Weight (kg) | n | 95 | 136 |
|  | Mean (SD) | 60.952 (20.6625) | 69.084 (16.7309) |
| BMI (kg/m<sup>2</sup>) | n | 89 | 136 |
|  | Mean (SD) | 22.007 (5.6779) | 24.019 (5.2279) |
| Systolic Blood Pressure (mmHg) | n | 82 | 136 |
|  | Mean (SD) | 121.4 (15.00) | 121.1 (13.54) |
| Diastolic Blood Pressure (mmHg) | n | 82 | 136 |
|  | Mean (SD) | 73.2 (10.53) | 75.3 (8.65) |
| Heart Rate (beats/min) | n | 82 | 136 |
|  | Mean (SD) | 85.2 (15.40) | 79.8 (12.58) |
| ADL Total Score | n | 124 | 136 |
|  | Mean (SD) | 11.78 (5.937) | 12.51 (4.947) |
| GAA1 Repeat Length | n | 129 | 119 |
|  | Mean (SD) | 589.7 (245.50) | 720.9 (269.58) |
|  | ≥ 675, n (%) | 54 (41.9%) | 66 (55.5%) |
| GAA2 Repeat Length | n | 121 | 116 |
|  | Mean (SD) | 862.8 (232.38) | 727.6 (296.93) |

Only patients with available information are summarized for each parameter.
Abbreviations: ADL=Activities of Daily Living; BMI=body mass index; FA=Friedreich
ataxia; FARS=Friedreich ataxia rating scale; mFARS=modified Friedreich ataxia rating scale

### Propensity of matching

For the propensity score model used in this analysis, the diagnostics used to assess comparability of the FA-COMS and MOXIe Extension subjects indicated that the quality of the matches was good for all three populations (**Table 5**; **Supplementary Table 6).** There were some instances where diagnostics feel below the “acceptable” range [29] (PROC PSMATCH documentation) for the comparison of the variances of the residuals for covariates in the two groups. This was true for the ratio of the variances of the residuals for age and age of onset covariates which were more variable than other covariates in the model (**Table 3; Supplementary Table 6**). Taken together, the diagnostic results show a high quality of matching for the Primary Pooled Population, Primary Placebo-Omav Population, and Primary Omav-Omav Population.

**Table 5:** Propensity Score Diagnostic Results.

| Diagnostic | Criteria<br>for Good or<br>Acceptable<br>Match<sup>a</sup> | Pooled<br>(Match 1) | Placebo-Omav<br>(Match 2) | Omav-Omav<br>(Match 3) |
| --- | --- | --- | --- | --- |
|  |  | Score | Score | Score |
| Standardized Difference of the Means of the Propensity Score | <0.5 | 0.0055 | 0.0090 | 0.0012 |
| Standardized Difference of the Means of Covariates |  |  |  |  |
| Sex | <0.5 | 0 | 0 | 0 |
| Baseline Gait | <0.5 | 0.0672 | 0.0802 | 0.0325 |
| Baseline mFARS | <0.5 | 0.0826 | 0.1103 | 0.0828 |
| Age at baseline | <0.5 | 0.0375 | 0.0902 | 0.1357 |
| Age at FRDA Onset | <0.5 | 0.0292 | 0.0645 | 0.0424 |
| Ratio of the Variances of the Propensity Score | Close to 1;<br>>0.8 and<br><1.25 | 1.0243 | 1.0411 | 0.9974 |
|  |  | Score | Score | Score |
| Ratio of the Variances of the Residuals for Covariates |  |  |  |  |
| Sex | 0.5 to 2 | 0.9999 | 1.0044 | 0.9993 |
| Baseline Gait | 0.5 to 2 | 0.5751 | 0.5022 | 0.5599 |
| Baseline mFARS | 0.5 to 2 | 0.6068 | 0.4986 | 0.5479 |
| Age at baseline | 0.5 to 2 | 0.3428 | 0.3305 | 0.2005 |
| Age at FRDA Onset | 0.5 to 2 | 0.3194 | 0.2852 | 0.4325 |

Criteria for a “good” match shown for standardized difference of the means of the propensity score, standardized difference of the means of the propensity score for each covariate, and ratio of the variances of the propensity score. Criteria for an “acceptable” match shown for the ratio of the variances of the residuals for each covariate.

### Efficacy Results

Results for the primary endpoint, change from baseline in mFARS score at Year 3, differed significantly between patients receiving omaveloxolone in MOXIe Extension and the untreated matched FA-COMS patients. After 3 years, in the Pooled Primary Population, matched FA-COMS patients progressed 6.6 mFARS points whereas patients treated with omaveloxolone in MOXIe Extension progressed 3.0 points (difference= -3.6 points; nominal p=0.0001) (**Table 6A**); thus, progression in mFARS was reduced by 55% in the omav treatment group compared to the matched NH group.. Analysis of the primary endpoint in the Primary Placebo-Omav and Primary Omav-Omav populations yielded similar results, with nominal p-values of < 0.05 for the treatment difference and slowing of progression in mFARS of >50% (**Table 6B**). In the Primary Placebo-Omav Population, containing treatment-naïve patients at Extension baseline, progression in mFARS was slowed by 56% compared to NH controls. In the Primary Omav-Omav Population, in which Extension patients had previously received 48 weeks of omaveloxolone treatment in Part 2, progression in mFARS was slowed by 61% in MOXIe Extension patients compared to FA-COMS patients, suggesting that such patients continue to benefit from omaveloxolone treatment (**Table 6B**).

**Table 6:**
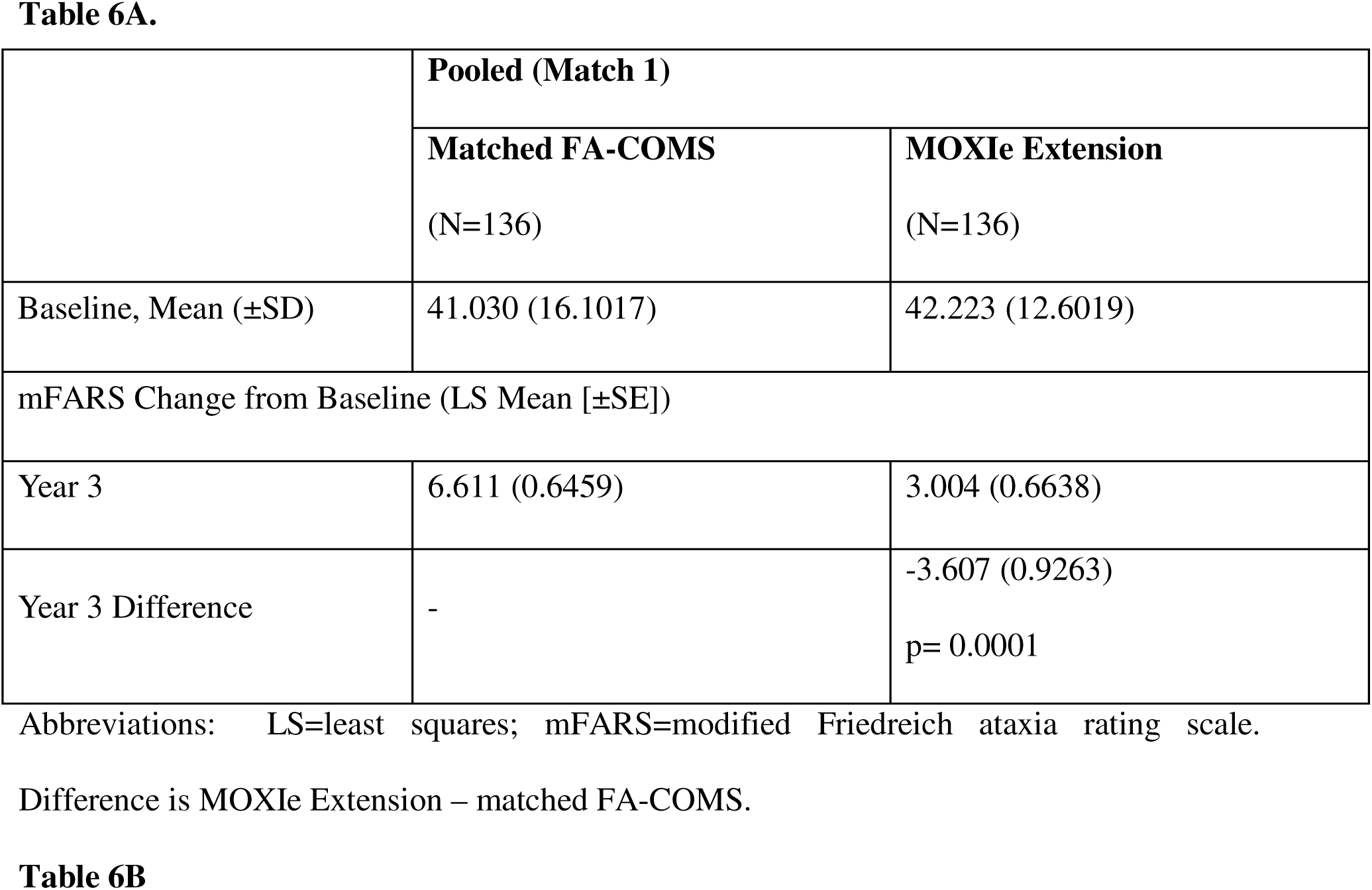
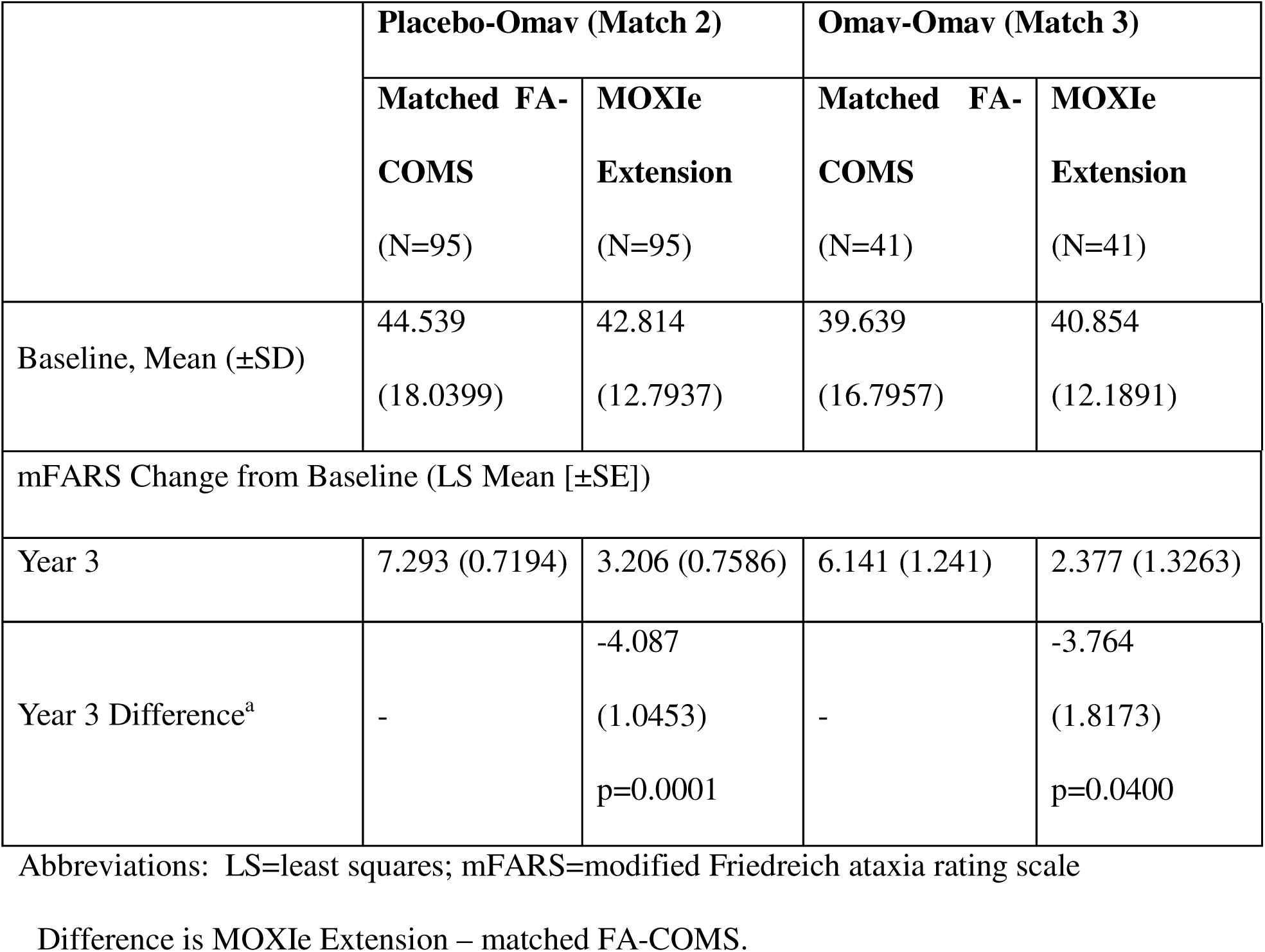
Analysis of Primary Outcome Measure

Results of the secondary endpoints, change from baseline in mFARS score at Year 1 and Year 2, also favor omaveloxolone in the Primary Pooled Population (**Table 7**; **Figure 3A**). At each year, Extension patients experienced a smaller change from baseline in mFARS than the matched FA-COMS patients. These distinct trajectories over 3 years show consistent separation between the 2 groups and yield nominal p-values less than 0.05 for all comparisons. In the Primary Placebo-Omav Population, the treatment effect favored MOXIe Extension at Year 1 and Year 2. MOXIe Extension patients experienced an improvement (i.e., a decrease) from baseline in mFARS at Year 1 and the treatment difference was -2.8 mFARS points (nominal p=0.0035) (**Table 7B**; **Figure 3B**). In the Primary Omav-Omav Population, the treatment effect favored the Extension at Year 1 and Year 2. A smaller difference between treatment groups was observed at Year 1 in the Omav-Omav Population than in the Placebo-Omav Population. Extension patients in the Omav-Omav Population did not experience an improvement from baseline, likely because they were in their second year of treatment with active drug. The treatment effect favored the Extension at each visit and consistently increased over time in this population (**Table 7C**; **Figure 3C**)

**Figure 3:**
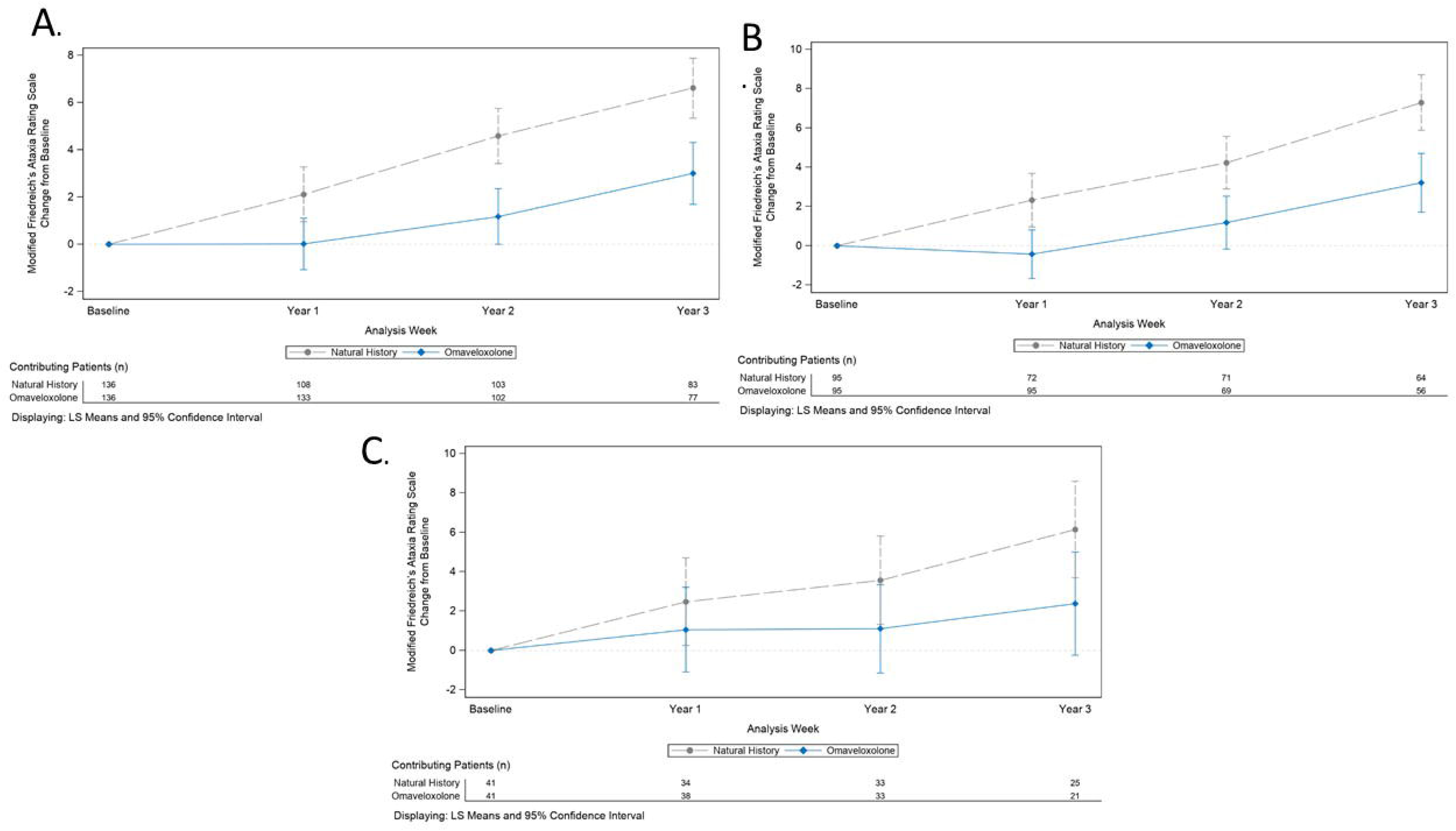
Change in mFARS from baseline of time. A. Primary Pooled Population B. Primary Placebo-Omav Population C. Primary Omav-Omav Population

**Table 7:**
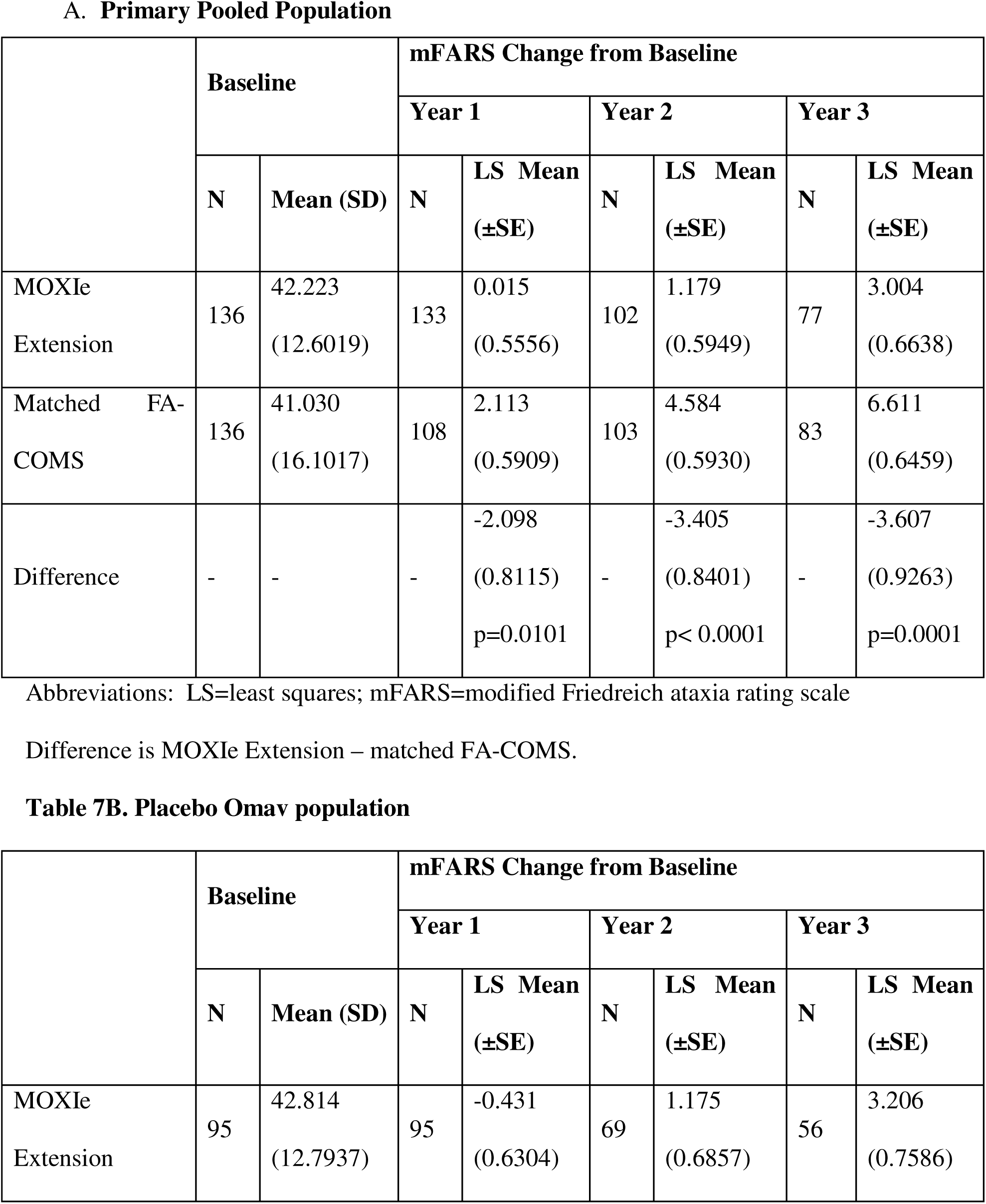
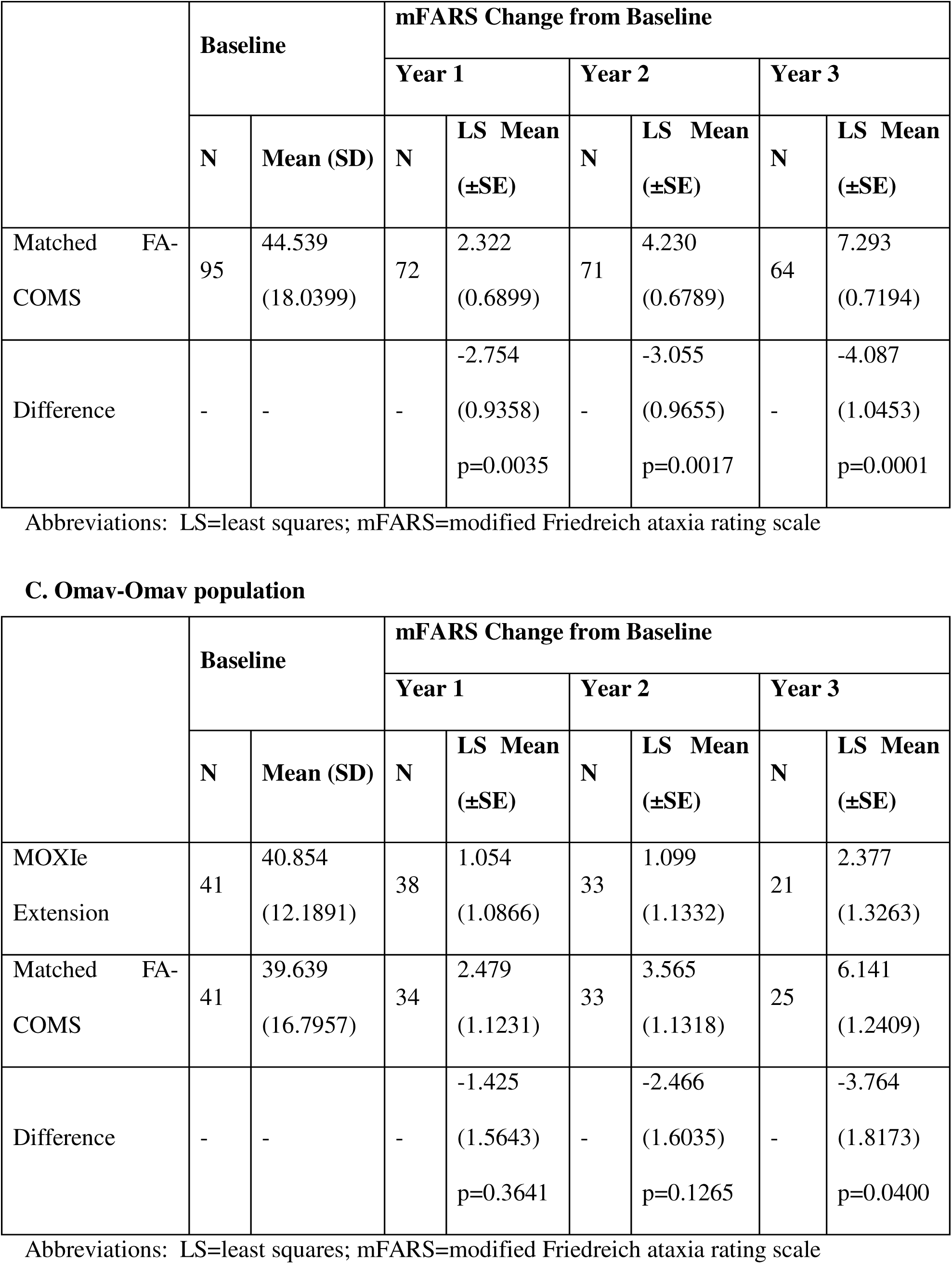
Change in mFARS over 3 years

### Sensitivity Analyses

Results of the primary and secondary endpoints were similar in the sensitivity populations, in which the pool of FA-COMS patients available for matching for each Extension set was further restricted to patients who had a baseline mFARS value and baseline age within the range observed in the MOXIe Extension baseline. At all timepoints, and in all three sensitivity populations (comprised of three separate sets of rematched patients), results favored omaveloxolone (Extension patients). At Year 3, the 3 sensitivity populations showed a treatment benefit of -2.4, -3.2, and -4.7 mFARS points for the Sensitivity Pooled, Sensitivity Placebo-Omav, and Sensitivity Omav-Omav populations, respectively, all with nominal p-values of <0.05 (**Supplementary Table 7**).

## DISCUSSION

The results of the present post-hoc analyses demonstrate that treatment with omaveloxolone provides a clinically meaningful slowing of FRDA progression over a 3-year period compared to untreated, propensity score-matched FA-COMS external controls. Data from Parts 1 and 2 of MOXIe were not included in this propensity-matched analysis. After 3 years in the largest analysis population, the Primary Pooled Population (n=272; 136 from FA-COMS and 136 from the MOXIe Extension), matched FA-COMS patients had progressed 6.6 mFARS points whereas patients treated with omaveloxolone in the MOXIe Extension progressed 3.0 points (difference = -3.6, nominal p=0.0001); thus, progression in mFARS was slowed by 55% with omaveloxolone treatment relative to natural history controls. Clinical trials in neurodegenerative diseases are typically powered to detect a 50% slowing of progression in 1 year [21], showing the context of the potential benefit here relative to other studies. During treatment in the MOXIe Extension, omaveloxolone treatment in the primary pooled population slowed progression by >50% at each year over 3 years compared to the corresponding FA-COMS external control group, indicating a benefit that persisted and accrued over the course of 3 years. Subsets of MOXIe Extension patients that differed in prior treatment status also had slowing of disease progression when matched to FA-COMS controls. All of the analysis populations suggested a benefit of omaveloxolone as did sensitivity analyses at all points during the 3-year duration of the study. In the MOXIe Part 2, omaveloxolone-treated patients experienced a reduction (i.e., an improvement) in mFARS score relative to the placebo group, revealing a benefit of omaveloxolone [19–20]. Together, the present results show a meaningful slowing of FRDA progression with omaveloxolone over a period of 3 years and further reinforce the findings of clinical benefit from MOXIe study Part 2 [19–20]. Thus, the benefit of omaveloxolone demonstrated across multiple analysis populations separately matched to FA-COMS patients and the comparability of these results to Part 2 results provides supportive evidence of for use of omaveloxolone in FRDA.

While there have been many attempts to use natural history studies as controls in clinical trials, the systematic design of such approaches is difficult. Analyses based on external controls are potentially limited by lack of randomization, which can introduce bias; however, ICH E10 guidelines advise that bias can be minimized when there is detailed patient information collected and when patients are as similar as possible, including in their demographic characteristics and baseline status, between interventional groups and natural history controls. The FA-COMS natural history study presents a unique opportunity for identification of suitable external control groups. The significant overlap in trial sites and investigators (some of whom helped develop FRDA standard of care guidelines), ensures a similar approach to overall patient management for FRDA and comorbid conditions [30]. In addition, there are no approved or effective disease modifying therapies for FRDA, minimizing confounding in the comparison of groups from 2 different studies. The overlap in trial sites and the reliance on the same investigator to provide training on mFARS assessments further contributed to consistency in methodology and scoring of mFARS in patients originating from both studies.

Use of propensity score matching provides further rigor in identifying an appropriate external control group that aligns with ICH E10 guidance. Using this approach, the matched FA-COMS group in the Primary Pooled Population was highly comparable for demographics and baseline characteristics to the MOXIe Extension patients, including the characteristics for determining the propensity scores. Similarly, in the Sensitivity Pooled Population, in which the matched set for the propensity score analysis was selected from a more restricted pool of patients, demographics and baseline characteristics were highly comparable. Additionally, propensity score matching diagnostic results demonstrated that the propensity score matching was good (i.e., < 0. 5) for the standardized difference of the means of the propensity score overall and for each covariate [24], good (i.e., close to 1; >0.8 and <1.25) for the ratio of the variances of the propensity score, and acceptable for the ratio of the variances of the residuals for covariates (0.5 to 2) for most covariates for all analysis populations. Although the model used for propensity score matching included multiple covariates that were considered prognostic for FRDA progression, the model may have been limited by not also having GAA1 repeat length or pes cavus as covariates. These were not included in the model due to lack of availability of GAA1 in all patients and differences in the method of evaluation of pes cavus between studies. However, baseline characteristics for mean GAA1 repeat length in both FA-COMS and MOXIe Extension patients were in a range in which the consequence of longer GAA1 repeat length on disease severity has reached a relative ceiling effect [25–27]. Furthermore, if it were significant, the slightly longer GAA1 length in MOXIe subjects would bias the results toward the null (as longer GAA1 lengths predict faster progression). While pes cavus might alter the response to treatment [19], it does not clearly influence the speed of disease progression.

Another potential limitation in the cross-study comparison was that scheduled assessments for mFARS differed for the 2 groups, with annual assessments in the FA-COMS study and biannual assessments in MOXIe Extension. Accordingly, analysis windows were defined using annual visits to align the MOXIe Extension schedule with the FA-COMS schedule. The MMRM analysis model using annual visits aligns with the MOXIe Part 2 primary analysis method, is a common model frequently used by the FDA, and does not rely on a linearity assumption. While the annual visits align with the FA-COMS assessment schedule, a limitation of the MMRM analysis using annual visits is not using all mFARS assessments from the every 24-week assessment schedule in MOXIe Extension.

In conclusion, although there are limitations to this cross-study analysis, the approach leads to readily interpretable results on the potential benefit of omaveloxolone in FRDA. The FA-COMS cohort identified by propensity score matching is highly comparable to the MOXIe Extension patients for both baseline characteristics and standard of care; therefore, the observed difference in disease progression (as assessed by mFARS) can be attributed to omaveloxolone treatment. Thus, such propensity-matched analysis data from the FA-COMS natural history study compared to MOXIe Extension provide evidence for the use of omaveloxolone for the treatment of FRDA and demonstrates the value and methodology for direct utilization of natural history data in clinical trials in rare diseases.

## Supporting information

Consort checklist

## Data Availability

All files from FACOMS data are held in a public repository at cPATH. (https://c-path.org/programs/rdca-dap/working-group/fa-icd/). Data from the MOXIe extension will not be available.

https://c-path.org/programs/dcc/projects/friedreichs-ataxia/

---

**Supplementary Table 1:**
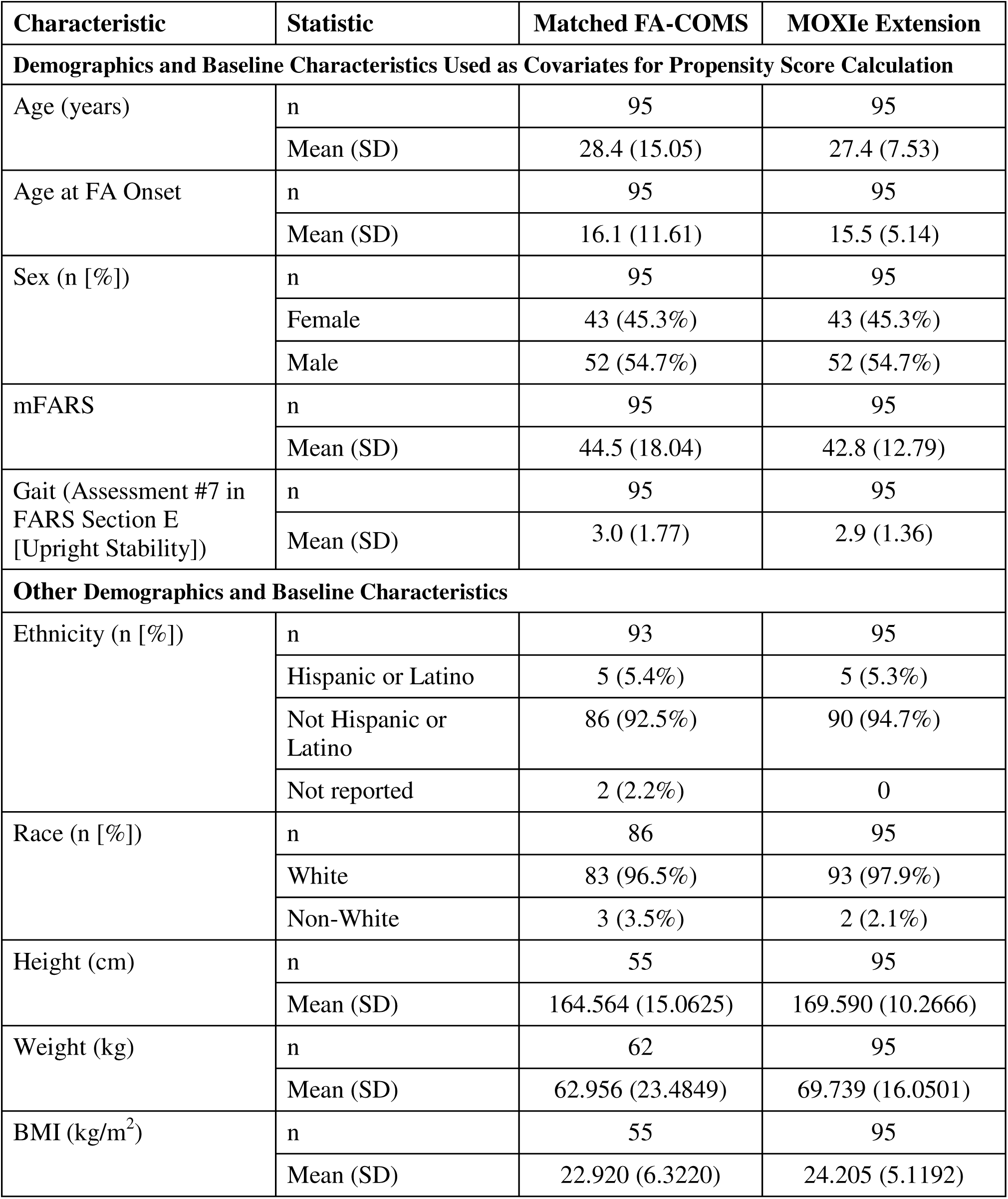
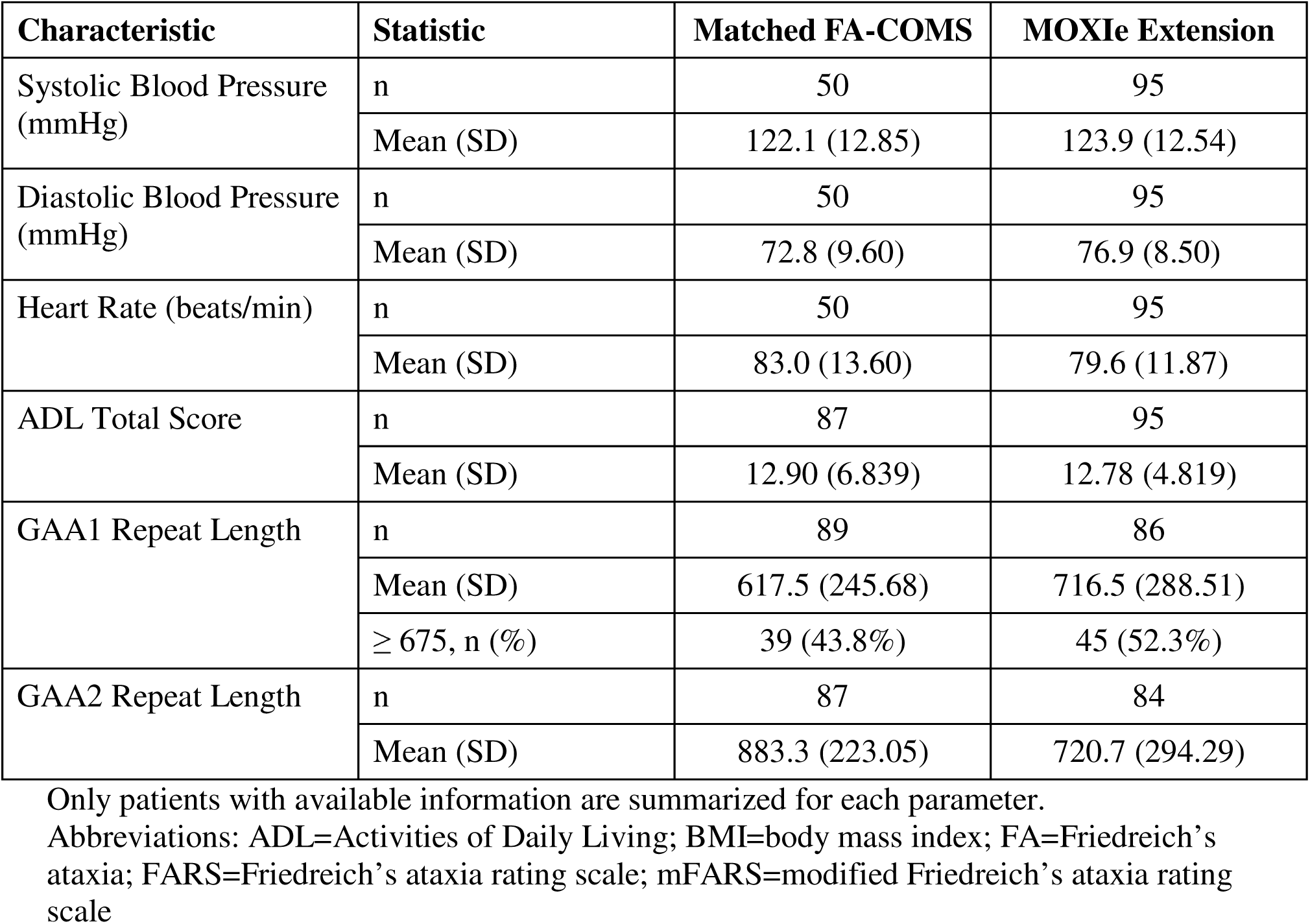
Demographics and Baseline Characteristics (Primary Placebo-Omav Population)

**Supplementary Table 2:**
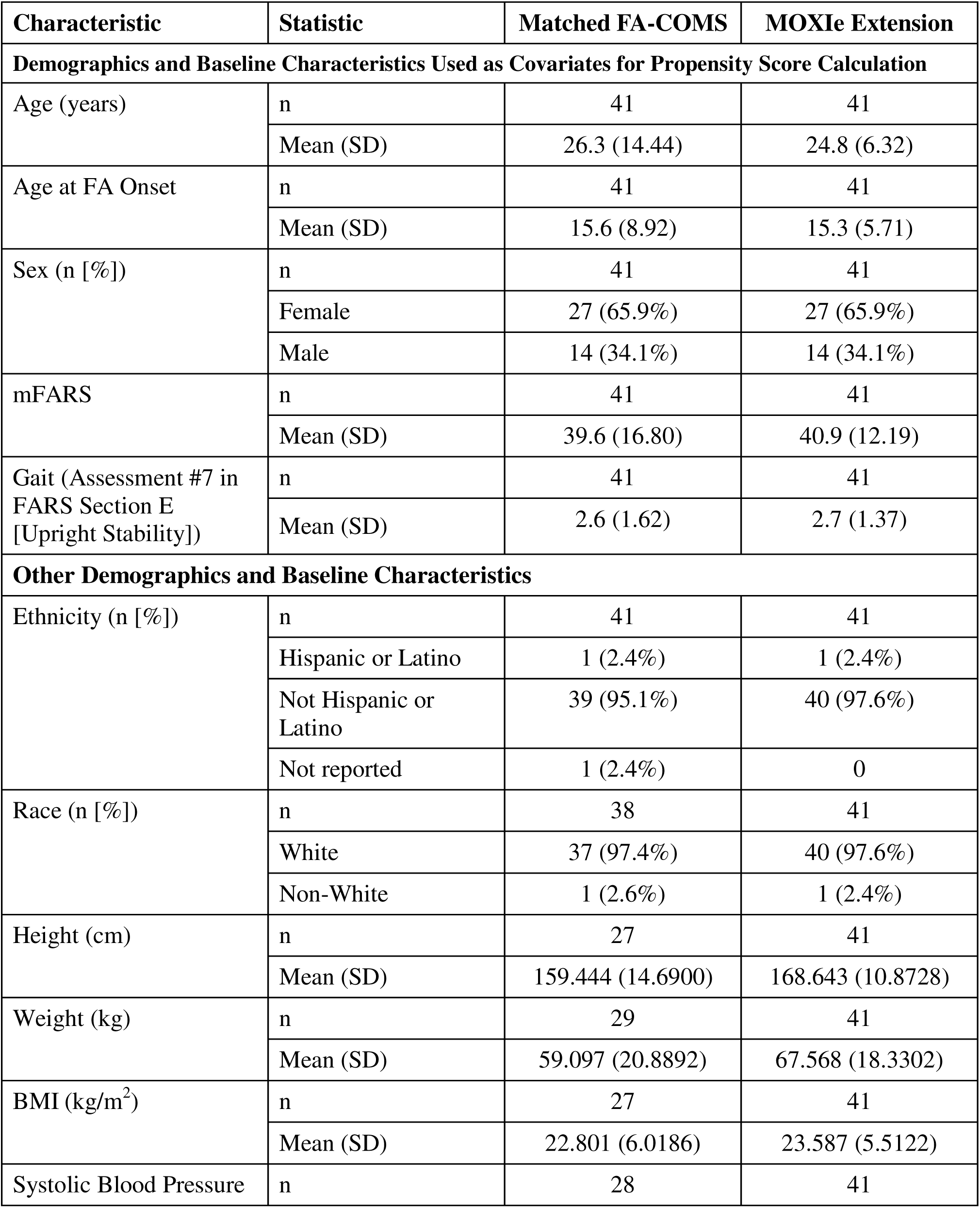
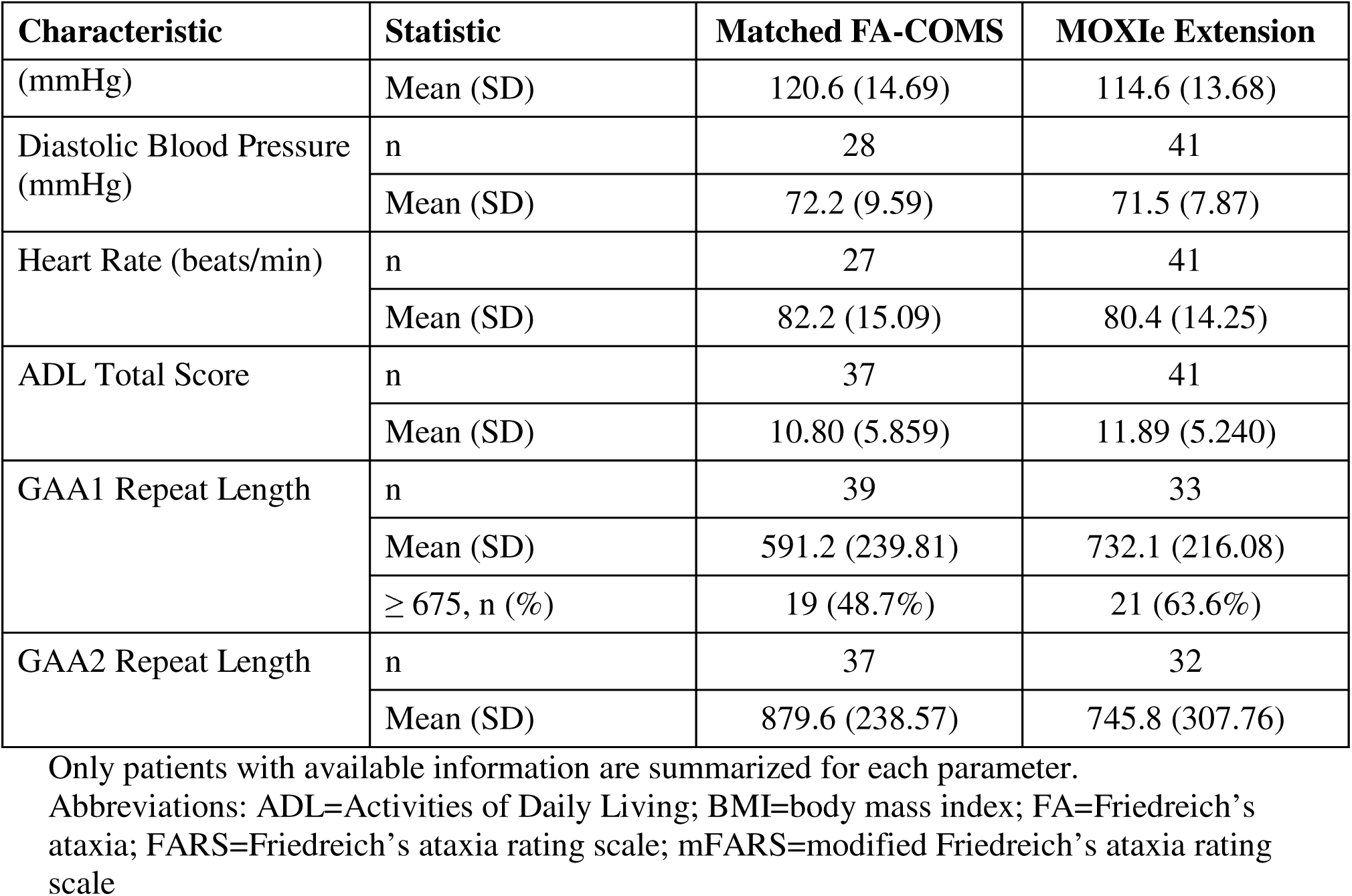
Demographics and Baseline Characteristics (Primary Omav-Omav Population)

**Supplementary Table 3:** Demographics and Baseline Characteristics (Sensitivity Pooled Population)

| Characteristic | Statistic | Matched FA-COMS | MOXIe Extension |
| --- | --- | --- | --- |
| **Demographics and Baseline Characteristics Used as Covariates for Propensity Score Calculation** |  |  |  |
| Age (years) | n | 133 | 133 |
|  | Mean (SD) | 26.5 (7.69) | 26.3 (7.07) |
| Age at FA Onset | n | 133 | 133 |
|  | Mean (SD) | 15.2 (5.52) | 15.2 (4.92) |
| Sex (n [%]) | n | 133 | 133 |
|  | Female | 67 (50.4%) | 67 (50.4%) |
|  | Male | 66 (49.6%) | 66 (49.6%) |
| mFARS | n | 133 | 133 |
|  | Mean (SD) | 43.8 (16.42) | 42.5 (12.60) |
| Gait (Assessment #7 in FARS Section E [Upright Stability]) | n | 133 | 133 |
|  | Mean (SD) | 2.9 (1.69) | 2.8 (1.37) |
| **Other Demographics and Baseline Characteristics** |  |  |  |
| Ethnicity (n [%]) | n | 132 | 133 |
|  | Hispanic or Latino | 2 (1.5%) | 5 (3.8%) |
|  | Not Hispanic or Latino | 126 (95.5%) | 128 (96.2%) |
|  | Not reported | 4 (3.0%) | 0 |
| Race (n [%]) | n | 126 | 133 |
|  | White | 122 (96.8%) | 130 (97.7%) |
|  | Non-White | 4 (3.2%) | 3 (2.3%) |
| Height (cm) | n | 77 | 133 |
|  | Mean (SD) | 171.013 (9.8962) | 169.344 (10.4826) |
| Weight (kg) | n | 85 | 133 |
|  | Mean (SD) | 66.815 (14.0819) | 68.836 (16.7214) |
| BMI (kg/m<sup>2</sup>) | n | 75 | 133 |
|  | Mean (SD) | 22.877 (4.2761) | 23.923 (5.2332) |
| Systolic Blood Pressure | n | 66 | 133 |

| **Characteristic** | **Statistic** | **Matched FA-COMS** | **MOXIe Extension** |
| --- | --- | --- | --- |
| (mmHg) | Mean (SD) | 123.4 (11.77) | 121.1 (13.66) |
| Diastolic Blood Pressure (mmHg) | n | 66 | 133 |
|  | Mean (SD) | 73.5 (8.87) | 75.2 (8.69) |
| Heart Rate (beats/min) | n | 66 | 133 |
|  | Mean (SD) | 80.9 (14.93) | 79.9 (12.62) |
| ADL Total Score | n | 125 | 133 |
|  | Mean (SD) | 12.34 (6.265) | 12.55 (4.886) |
| GAA1 Repeat Length | n | 127 | 116 |
|  | Mean (SD) | 573.5 (198.35) | 721.2 (270.97) |
|  | ≥ 675, n (%) | 41 (32.3%) | 64 (55.2%) |
| GAA2 Repeat Length | n | 122 | 113 |
|  | Mean (SD) | 869.9 (244.80) | 738.9 (290.46) |

Only patients with available information are summarized for each parameter.
Abbreviations: ADL=Activities of Daily Living; BMI=body mass index; FA=Friedreich
ataxia; FARS=Friedreich ataxia rating scale; mFARS=modified Friedreich ataxia rating scale

**Supplementary Table 4:**
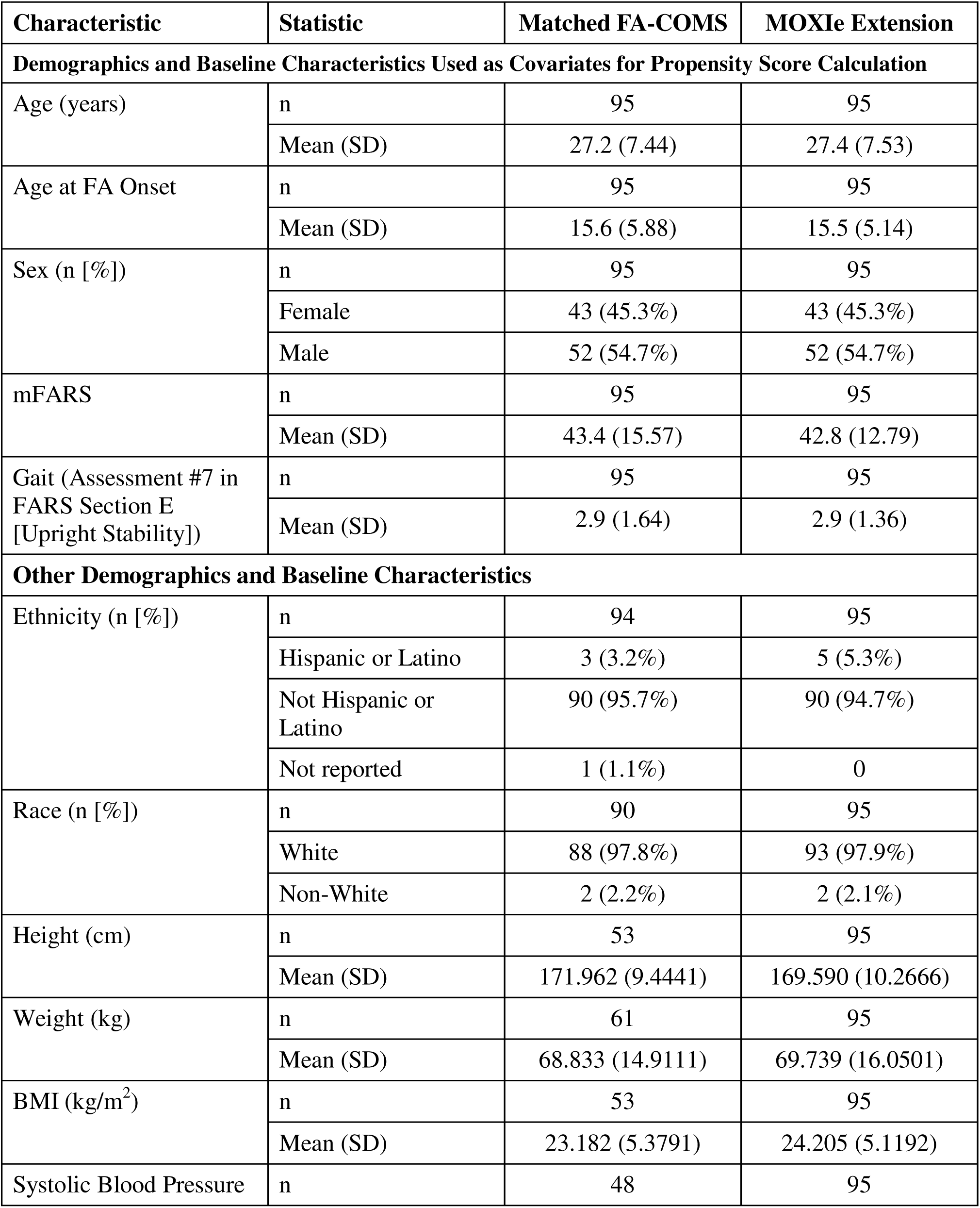
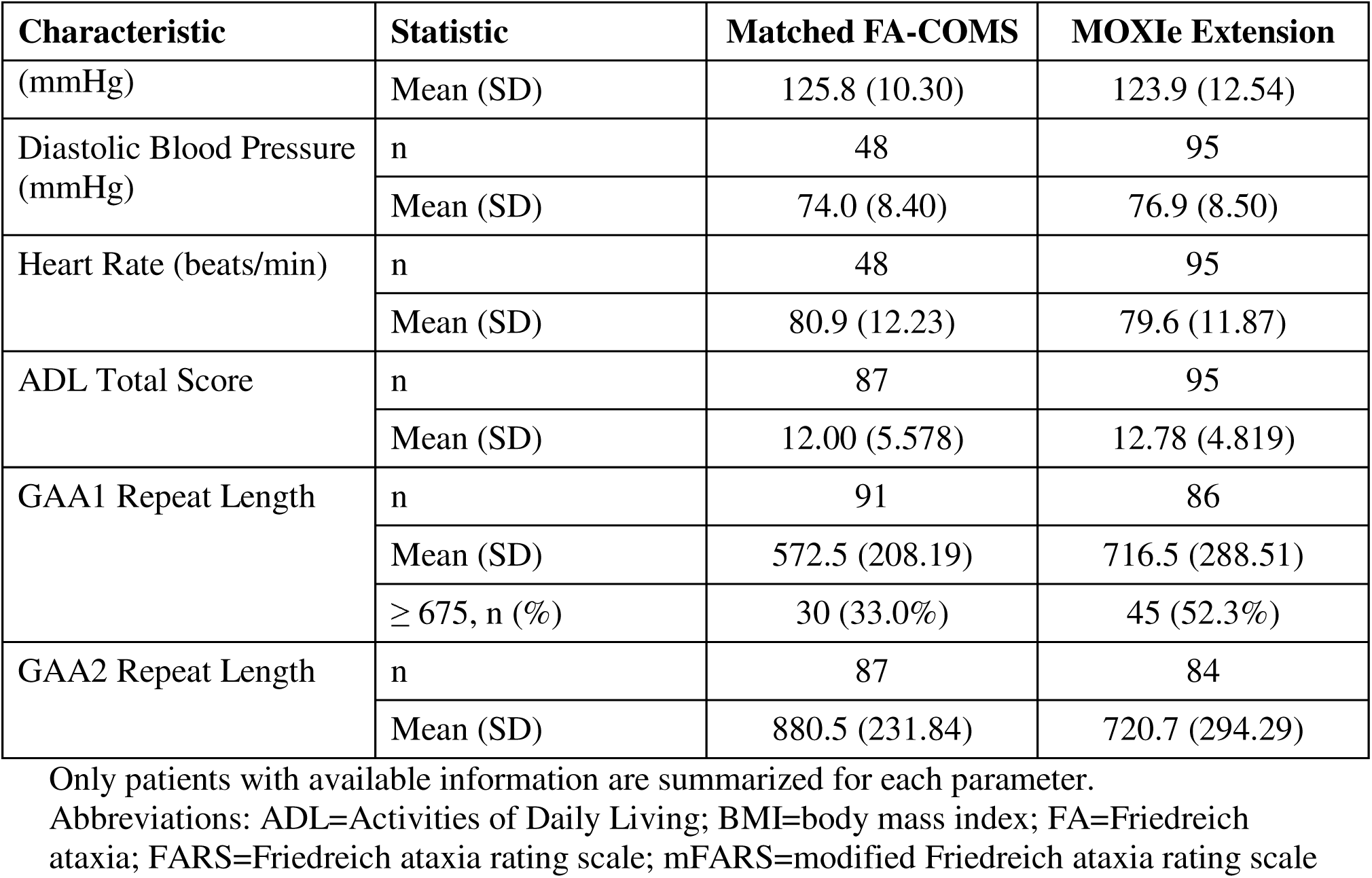
Demographics and Baseline Characteristics (Sensitivity Placebo-Omav Population)

**Supplementary Table 5:** Demographics and Baseline Characteristics (Sensitivity Omav-Omav Population)

| Characteristic | Statistic | Matched FA-COMS | MOXIe Extension |
| --- | --- | --- | --- |
| **Demographics and Baseline Characteristics Used as Covariates for Propensity Score Calculation** |  |  |  |
| Age (years) | n | 40 | 40 |
|  | Mean (SD) | 24.5 (6.77) | 24.5 (6.08) |
| Age at FA Onset | n | 40 | 40 |
|  | Mean (SD) | 14.9 (4.73) | 14.8 (4.70) |
| Sex (n [%]) | n | 40 | 40 |
|  | Female | 26 (65.0%) | 26 (65.0%) |
|  | Male | 14 (35.0%) | 14 (35.0%) |
| mFARS | n | 40 | 40 |
|  | Mean (SD) | 42.6 (16.26) | 41.2 (12.17) |
| Gait (Assessment #7 in FARS Section E [Upright Stability]) | n | 40 | 40 |
|  | Mean (SD) | 2.8 (1.71) | 2.7 (1.38) |
| **Other Demographics and Baseline Characteristics** |  |  |  |
| Ethnicity (n [%]) | n | 38 | 40 |
|  | Hispanic or Latino | 2 (5.3%) | 1 (2.5%) |
|  | Not Hispanic or Latino | 35 (92.1%) | 39 (97.5%) |
|  | Not reported | 1 (2.6%) | 0 |
| Race (n [%]) | n | 35 | 40 |
|  | White | 35 (100%) | 39 (97.5%) |
|  | Non-White | 0 | 1 (2.5%) |
| Height (cm) | n | 27 | 40 |
|  | Mean (SD) | 169.444 (9.2542) | 168.446 (10.9368) |
| Weight (kg) | n | 27 | 40 |
|  | Mean (SD) | 61.848 (14.5368) | 66.924 (18.0870) |
| BMI (kg/m<sup>2</sup>) | n | 27 | 40 |
|  | Mean (SD) | 21.601 (5.3272) | 23.432 (5.4908) |
| Systolic Blood Pressure (mmHg) | n | 23 | 40 |
|  | Mean (SD) | 121.3 (12.53) | 114.2 (13.60) |
| Diastolic Blood Pressure (mmHg) | n | 23 | 40 |
|  | Mean (SD) | 73.9 (9.57) | 71.2 (7.62) |
| Heart Rate (beats/min) | n | 23 | 40 |
|  | Mean (SD) | 80.7 (15.65) | 80.7 (14.27) |
| ADL Total Score | n | 38 | 40 |
|  | Mean (SD) | 11.46 (6.400) | 12.14 (5.059) |
| GAA1 Repeat Length | n | 40 | 32 |
|  | Mean (SD) | 585.1 (200.94) | 725.8 (216.46) |
|  | ≥ 675, n (%) | 17 (42.5%) | 20 (62.5%) |
| GAA2 Repeat Length | n | 39 | 31 |
|  | Mean (SD) | 872.1 (245.93) | 758.1 (304.81) |

Only patients with available information are summarized for each parameter.
Abbreviations: ADL=Activities of Daily Living; BMI=body mass index; FA=Friedreich ataxia; FARS=Friedreich ataxia rating scale; mFARS=modified Friedreich ataxia rating scale

**Supplementary Table 6:**
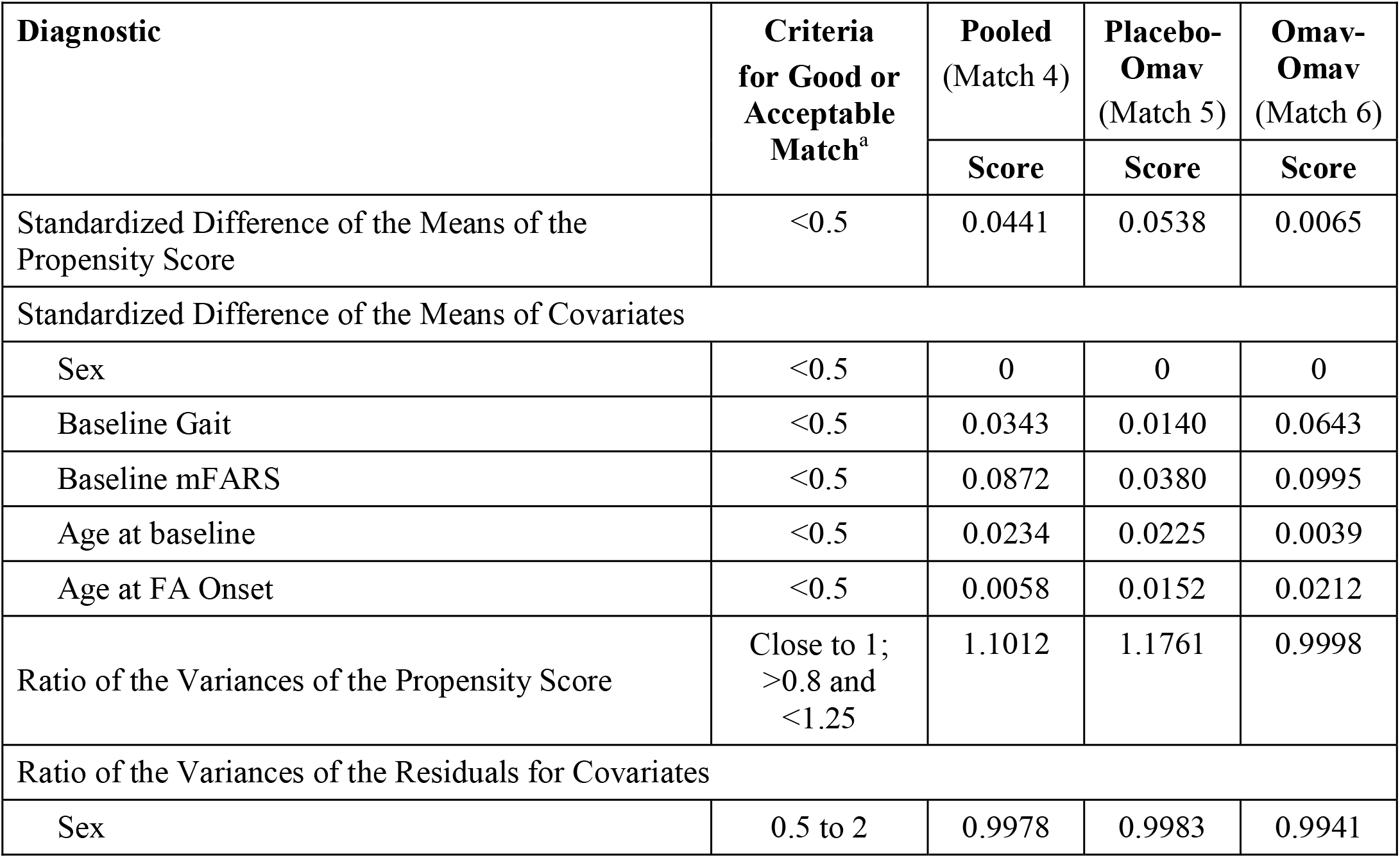
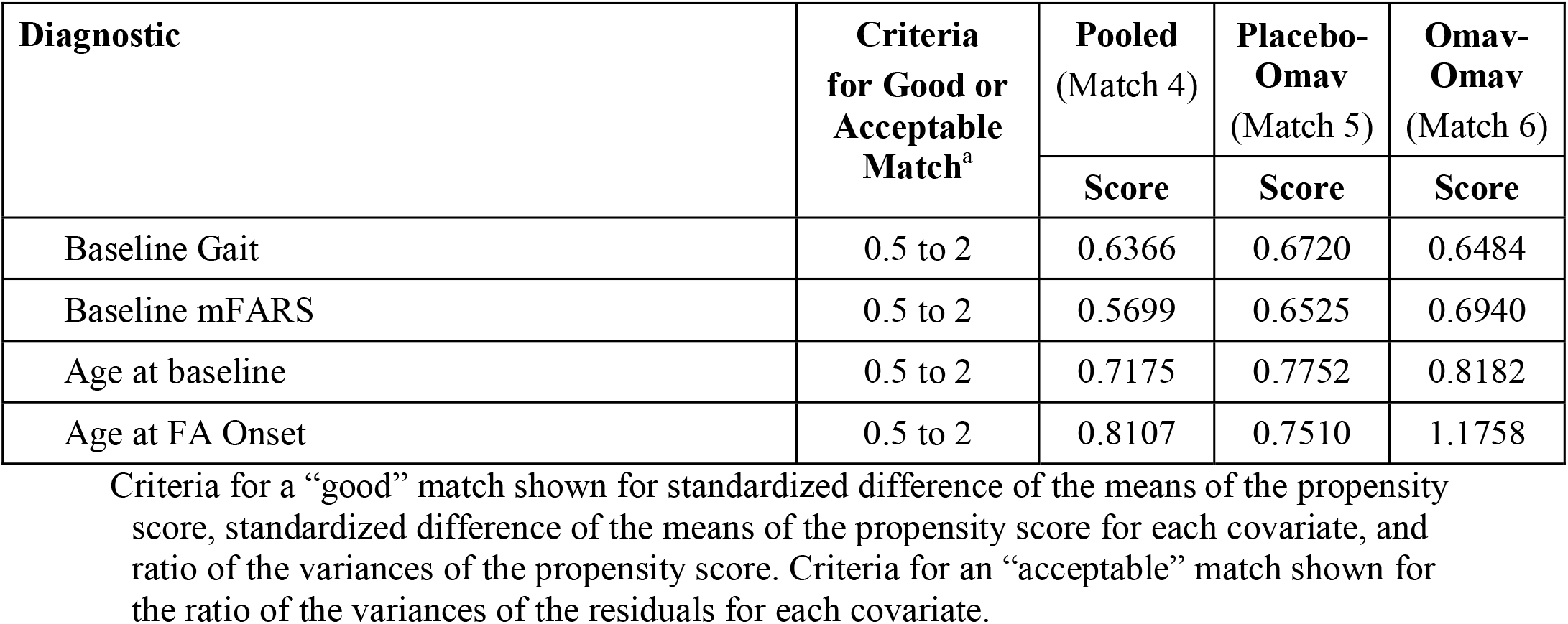
Propensity Score Diagnostic Results (Sensitivity Populations)

**Supplementary Table 7:**
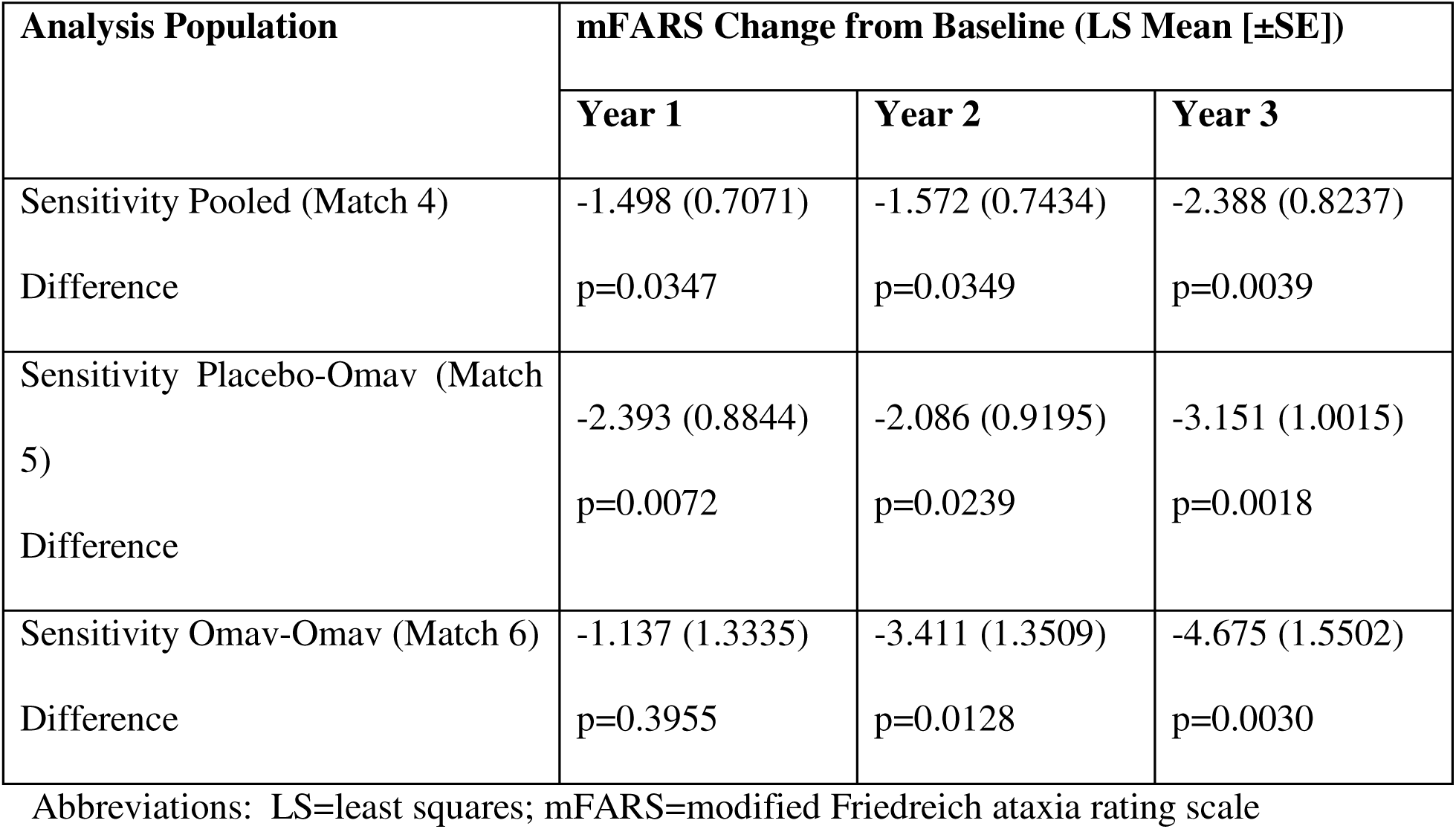
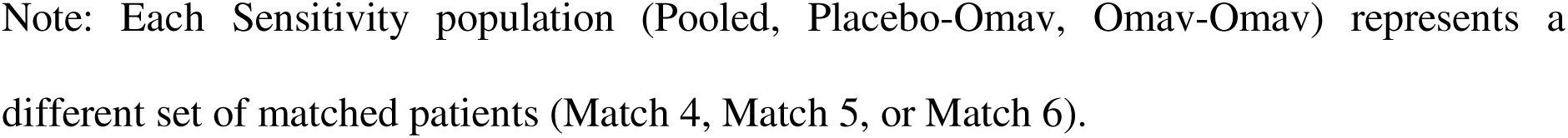
mFARS Change in Sensitivity Populations

| Analysis Population | mFARS Change from Baseline (LS Mean [±SE]) |  |  |
| --- | --- | --- | --- |
|  | Year 1 | Year 2 | Year 3 |
| Sensitivity Pooled (Match 4) | -1.498 (0.7071) | -1.572 (0.7434) | -2.388 (0.8237) |
| Difference | p=0.0347 | p=0.0349 | p=0.0039 |
| Sensitivity Placebo-Omav (Match 5) | -2.393 (0.8844) | -2.086 (0.9195) | -3.151 (1.0015) |
| Difference | p=0.0072 | p=0.0239 | p=0.0018 |
| Sensitivity Omav-Omav (Match 6) | -1.137 (1.3335) | -3.411 (1.3509) | -4.675 (1.5502) |
| Difference | p=0.3955 | p=0.0128 | p=0.0030 |

Abbreviations: LS=least squares; mFARS=modified Friedreich ataxia rating scale
Note: Each Sensitivity population (Pooled, Placebo-Omap, Omap-Omap) represents a different set of matched patients (Match 4, Match 5, or Match 6).

